# Women’s knowledge, attitudes and views of preconception health and intervention delivery methods: A cross-sectional survey

**DOI:** 10.1101/2022.05.26.22275637

**Authors:** Michael P Daly, James White, Julia Sanders, Ruth R Kipping

**Affiliations:** Bristol Medical School, University of Bristol, Bristol, UK; School of Medicine, Cardiff University, UK; School of Healthcare Sciences, Cardiff University, UK

**Keywords:** Preconception Care, Health Knowledge, Attitudes, Practice, Surveys and Questionnaires, Cross-Sectional Studies, Intervention development

## Abstract

**Background:** Several preconception exposures have been associated with adverse pregnancy, birth and postpartum outcomes. However, few studies have investigated women’s knowledge of and attitudes towards preconception health, and the acceptability of potential intervention methods.

**Methods:** Seven GP practices in the West of England posted questionnaires to 4,330 female patients aged 18 to 48 years. Without providing examples, we asked women to list maternal preconception exposures that might affect infant and maternal outcomes, and assessed their knowledge of nine literature-derived risk factors. Attitudes towards preconception health (interest, intentions, self-efficacy and perceived awareness and importance) and the acceptability of intervention delivery methods were also assessed. Multivariable multilevel regression examined participant characteristics associated with these outcomes.

**Results:** Of those who received questionnaires, 835 (19.3%) responded. Women were most aware of the preconception risk factors of diet (86.0%) and physical activity (79.2%). Few were aware of weight (40.1%), folic acid (32.9%), abuse (6.3%), advanced age (5.9%) and interpregnancy intervals (0.2%), and none mentioned interpregnancy weight change or excess iron intake. After adjusting for demographic and reproductive covariates, women aged 18-25-years (compared to 40-48-years) and nulligravid women were less aware of the benefit of preconception folic acid supplementation (adjusted odds ratios (aOR) for age: 4.30 [2.10-8.80], gravidity: aOR 2.48 [1.70-3.62]). Younger women were more interested in learning more about preconception health (aOR 0.37 [0.21-0.63]) but nulligravid women were less interested in this (aOR 1.79 [1.30-2.46]). Women with the lowest household incomes (versus the highest) were less aware of preconception weight as a risk factor (aOR: 3.11 [1.65-5.84]) and rated the importance of preconception health lower (aOR 3.38 [1.90-6.00]). The most acceptable information delivery channels were websites/apps (99.5%), printed healthcare materials (98.6%), family/partners (96.3%), schools (94.4%), television (91.9%), pregnancy tests (91.0%) and doctors, midwives and nurses (86.8-97.0%). Dentists (23.9%) and hairdressers/beauticians (18.1%) were the least acceptable.

**Conclusions:** Our findings demonstrate a need to promote awareness of preconception risk factors and motivation for preconception health changes, particularly amongst younger and nulligravid women and women with lower incomes. Interventions to improve preconception health should focus on communication from healthcare professionals, schools, family members, and digital media.

## Background

Adverse pregnancy, birth and postpartum outcomes have a substantial impact on morbidity and mortality. Worldwide, there are around 23 million miscarriages (1), 14.8 million live preterm births (2), 2.4 million neonatal deaths (3) and 295,000 maternal deaths due to pregnancy or childbirth complications each year (4). A recent umbrella review found high- and moderate-certainty evidence that maternal preconception folate supplementation, maternal body mass index (BMI), interpregnancy weight change and physical inactivity affect the risk of outcomes such as neural tube defects, pre-eclampsia and gestational diabetes (5). The guidelines and policies of national health organisations also highlight the importance of these factors in the preconception period (6-8). However, evidence suggests that less than half of women begin folate supplementation before pregnancy in countries such as England, Scotland and the United States (9, 10), one in two women of reproductive age are overweight or obese (10, 11) and, globally, 31.7% are not sufficiently active (12). Moreover, a number of these preconception risk factors have been associated with maternal characteristics such as age, ethnicity, educational attainment, socioeconomic status, reproductive history and pregnancy intentions (10, 13-16).

Improving women’s knowledge of and attitudes towards preconception health is considered crucial to improving child and maternal health (17). There are, however, limitations in the existing literature on these factors. A recent systematic review of studies that measured women’s knowledge of preconception health (18) found 18 of the 34 included studies were of low methodological quality, with almost half recruiting only student participants and most omitting preconception risk factors such as maternal BMI and physical inactivity. Studies also prompted participants by listing risk factors, which may lead to an overestimation of knowledge. The one study that used open-ended, free-text questions (19) to assess this knowledge only recruited university students (N = 299) and asked about lifestyle changes. These differences in sampling and question design have resulted in heterogenous knowledge estimates, with estimates ranging from 31% to 100% reported for women’s knowledge of recommendations to supplement with folic acid before pregnancy (15).

The last and only study to have assessed British women’s knowledge of and attitudes towards preconception health was published by Wallace and Hurwitz in 1998 (20). The authors found Asian ethnicity, foreign birth, and nulligravidity were associated with a lack of knowledge of preconception folate supplementation, recommended alcohol consumption limits, and rubella infection indications. The delivery of preconception care in primary was found to be acceptable to participants, but no other methods of intervention delivery were explored. The study also only recruited patients who attended nine general practices in an affluent London borough during the recruitment period, which may have resulted in an overestimation of knowledge. Moreover, only univariate statistical techniques were used, meaning covariates were not adjusted for and the reported findings may have been spurious.

Accordingly, our objectives were to assess: women’s knowledge of preconception exposures associated with risk of adverse pregnancy, birth and postpartum outcomes, their attitudes towards preconception health, and the acceptability of different delivery methods for preconception health interventions. We also explored whether these varied by demographic characteristics, reproductive history, and pregnancy intentions.

## Methods

The study received ethical approval from the South West – Frenchay Research Ethics Committee before its conduct. It is reported following STROBE guidelines (21).

### Study design

A cross-sectional survey design was used, informed by theories of behaviour (22). We assessed: women’s knowledge of preconception health risk factors and their intentions and self-efficacy in making positive preconception changes; their perceived awareness of preconception health risk factors and attitudes towards the importance of preconception health and learning more about it; and the acceptability of different methods of delivering interventions to improve preconception health.

### Setting

Seven general practices in the West of England were purposively sampled to maximise variation in the socioeconomic backgrounds of participants, through considering indices of deprivation for each prospective practice’s locality (23) and oversampling practices from relatively deprived areas. Practices sent eligible patients the questionnaire by post, between August 2020 and March 2021. Prepaid envelopes, an information sheet and study team contact details were also enclosed. The first page of the questionnaire informed recipients that to take part in the study, they had to read and tick each of four consent statements. Recipients were given the option of completing the questionnaire anonymously to reduce the risk of socially desirable responding (24). Reminder postcards were sent to all recipients one month after the questionnaire. Participants were given a £5 shopping voucher to thank them for taking part.

### Participants

Lists of women meeting the study’s inclusion criteria (Box 1) were compiled by practice administrators using standardised electronic patient database searches. General practitioners (GPs) then worked through these lists consecutively, consulting patient notes where further information was required, to remove patients according to the study’s exclusion criteria until the required numbers of eligible patients were identified. The eligibility criteria were informed by the literature and input from GPs based at non-participating practices.

The upper age threshold of 48 years was chosen to facilitate comparison with the findings of Wallace and Hurwitz’s (20) survey, and the lower threshold of 18 years was selected as the consulted GPs felt it would be unacceptable to send the questionnaire to non-adult participants. Eligible patients were required to have English as their main spoken language as many of the questionnaire’s items have not been validated for use in non-English speaking populations, and because funding was not available for the use of translators. The remaining criteria were developed to exclude patients who were likely to be distressed by a questionnaire relating to pregnancy and/or have difficulties providing informed consent.

**Box 1:**
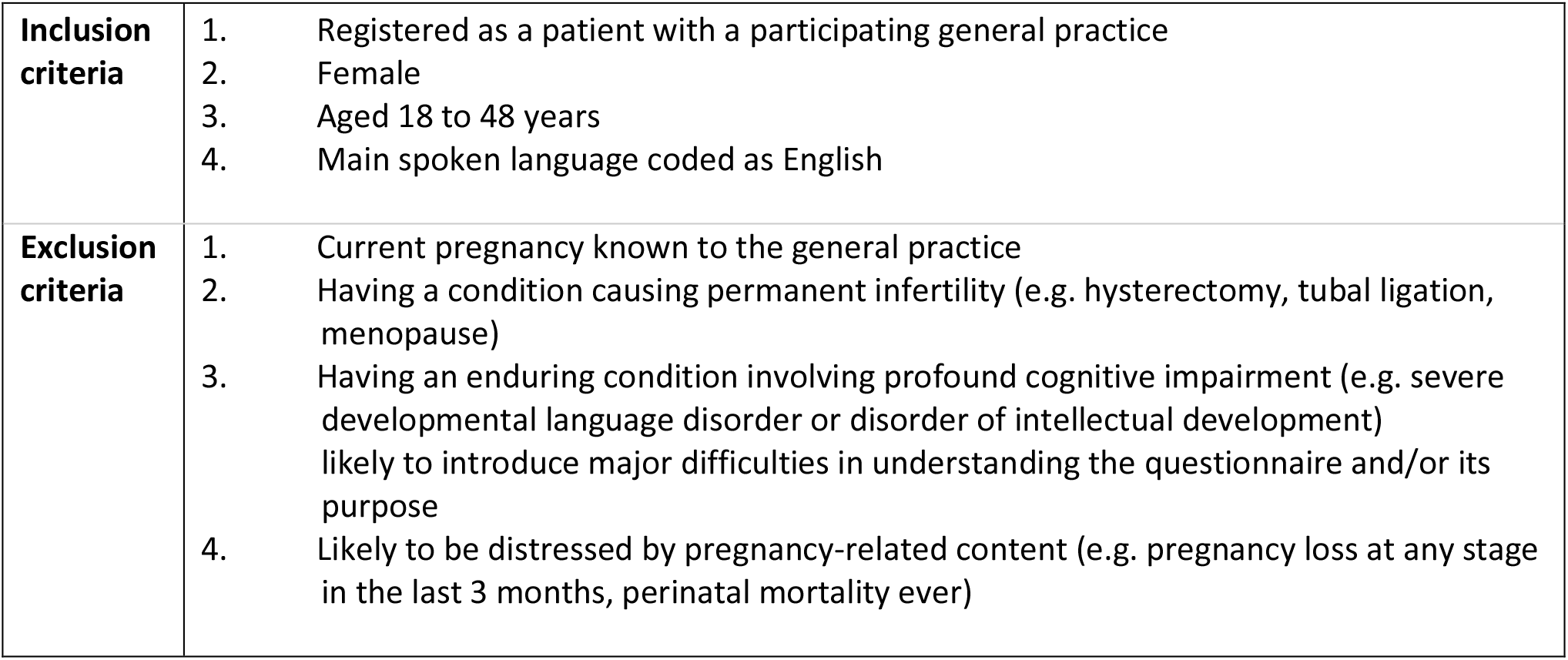
Study eligibility criteria

Box 1: Study eligibility criteria
|  |  |
| --- | --- |
| **Inclusion criteria** | 1. Registered as a patient with a participating general practice<br>2. Female<br>3. Aged 18 to 48 years<br>4. Main spoken language coded as English |
| **Exclusion criteria** | 1. Current pregnancy known to the general practice<br>2. Having a condition causing permanent infertility (e.g. hysterectomy, tubal ligation, menopause)<br>3. Having an enduring condition involving profound cognitive impairment (e.g. severe developmental language disorder or disorder of intellectual development) likely to introduce major difficulties in understanding the questionnaire and/or its purpose<br>4. Likely to be distressed by pregnancy-related content (e.g. pregnancy loss at any stage in the last 3 months, perinatal mortality ever) |

### Variables and measurement

Full information on the question stems, response options, sources, psychometric properties, and modifications made to the questionnaire’s featured items can be found in Additional file 1. The participant characteristic exposure variables were: age, ethnicity, and country of birth (25), educational attainment (26), and household income (27). Additionally, items relating to gravidity, previous live birth(s), adverse pregnancy outcomes, fertility issues, and pregnancy intentions were adapted from the third British national survey of sexual attitudes and lifestyles (Natsal-3) questionnaire (28). Exposure variables were also used in the multivariable analyses as covariates.

The outcome measures were assessed using relevant questionnaire items from survey studies featured in three literature reviews (13, 15, 18). The open-ended, free-text item used by Stern et al. (19) was used to assess knowledge of maternal preconception risk factors. However, while the original item asked participants to list only ‘changes in lifestyle’, our adapted version asked women to list maternal pre-pregnancy factors that could be done, started, continued, stopped or avoided, or that ‘relate to a woman’s life, circumstances or health’, which might affect pregnancy outcomes. The number of participants who listed each of nine preconception risk factors (Box 2), for which high-, moderate-or low-certainty evidence of an association with an adverse pregnancy, birth or postpartum outcome(s) was found in a recent umbrella review (5), was summed. All other preconception exposures named as risk factors by participants were also tallied.

Informed by the COM-B model (22), questions on women’s attitudes included: perceived awareness (29) and importance (30) of preconception health, interest in knowing more about preconception health (31), and preconceptional self-efficacy (32) and lifestyle change intentions (19). No existing items relating to the use and acceptability of potential intervention delivery channels were identified, so we asked participants to rate how comfortable they would be discussing preconception health and their pregnancy plans with various people, when they last had contact with each of these people, and how acceptable it would be for information about preconception health to be made available in different places and settings, using five- and seven-point Likert scales. These intervention delivery methods were derived from the literature and discussions within the study team (see Additional file 2).

**Box 2:**
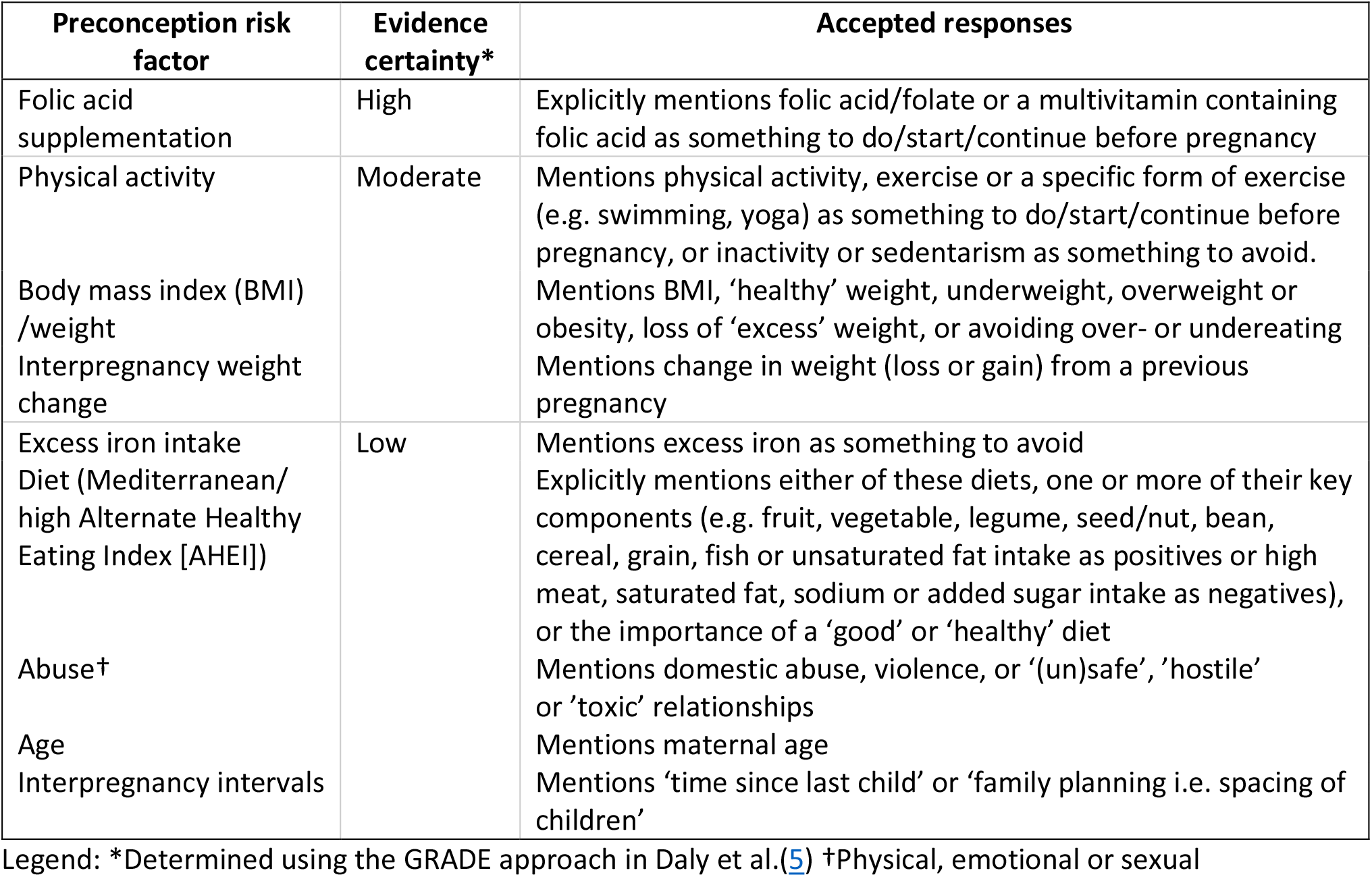
Preconception risk factors, with accepted participant responses

To examine content validity, the full questionnaire was presented to an external ‘expert panel’ as recommended by de Vet et al. (33), involving academics, a pregnancy charity representative and a public health lead, all of whom had expertise in preconception health specifically or child and maternal health generally. To establish face validity, twenty-one female members of the public aged 18-48-years were consulted to ensure the questionnaire’s phrasing and formatting were comprehensible and acceptable to the target population. Recurring departures from the intended interpretation were noted and commonly misunderstood terms and phrasings were either re-worded or explicitly defined.

### Study size and statistical methods

The target sample size was estimated as 770. This was calculated with the prevalence of the outcome variables as 50% in both the 18-29 and 30-48-years age groups, as recommended for instances where prevalence is unknown (34), and by inputting a precision of 5%, giving a 95% confidence interval of 45-55%.

Having a similar number of 18-29 and 30-48-year-olds was of interest as 29 years has been the national average maternal age at first birth for the past seven years (35). Questionnaires were sent in two mailouts, with response rates from the first mailout informing the number of questionnaires required in the subsequent mail-out to reach the target sample sizes in both age groups.

Prevalence values with 95% confidence intervals were calculated for all outcome variables. Univariable multilevel regression (individuals nested within GP practices) was performed for each exposure.

Multivariable multilevel regression models were then performed to explore whether these exposure variables were associated with the outcomes following adjustment for covariates. Covariates were informed by the literature (9, 11, 14, 15, 19, 20) and chosen *a priori*. Covariates that were very weakly correlated with the exposure (r<0.05) or showed a weak association with the outcome (*p*>0.05) in the univariable analyses were not included to avoid unnecessary adjustment, which can adversely affect estimate precision in logistic models (36). The covariates adjusted for in each analysis are presented in Additional file 3. Analyses excluded participants missing data for any of the included variables.

## Results

### Participant characteristics

Figure 1 shows the flow of study participants. 725 women were excluded, with the most common reasons being: likely to be distressed by pregnancy-related content (65.5%), current pregnancy known to the practice (17.9%), and enduring and profound cognitive impairment (11.7%). 4,330 questionnaires were sent and 835 (19.3%) were returned.

**Figure 1:**
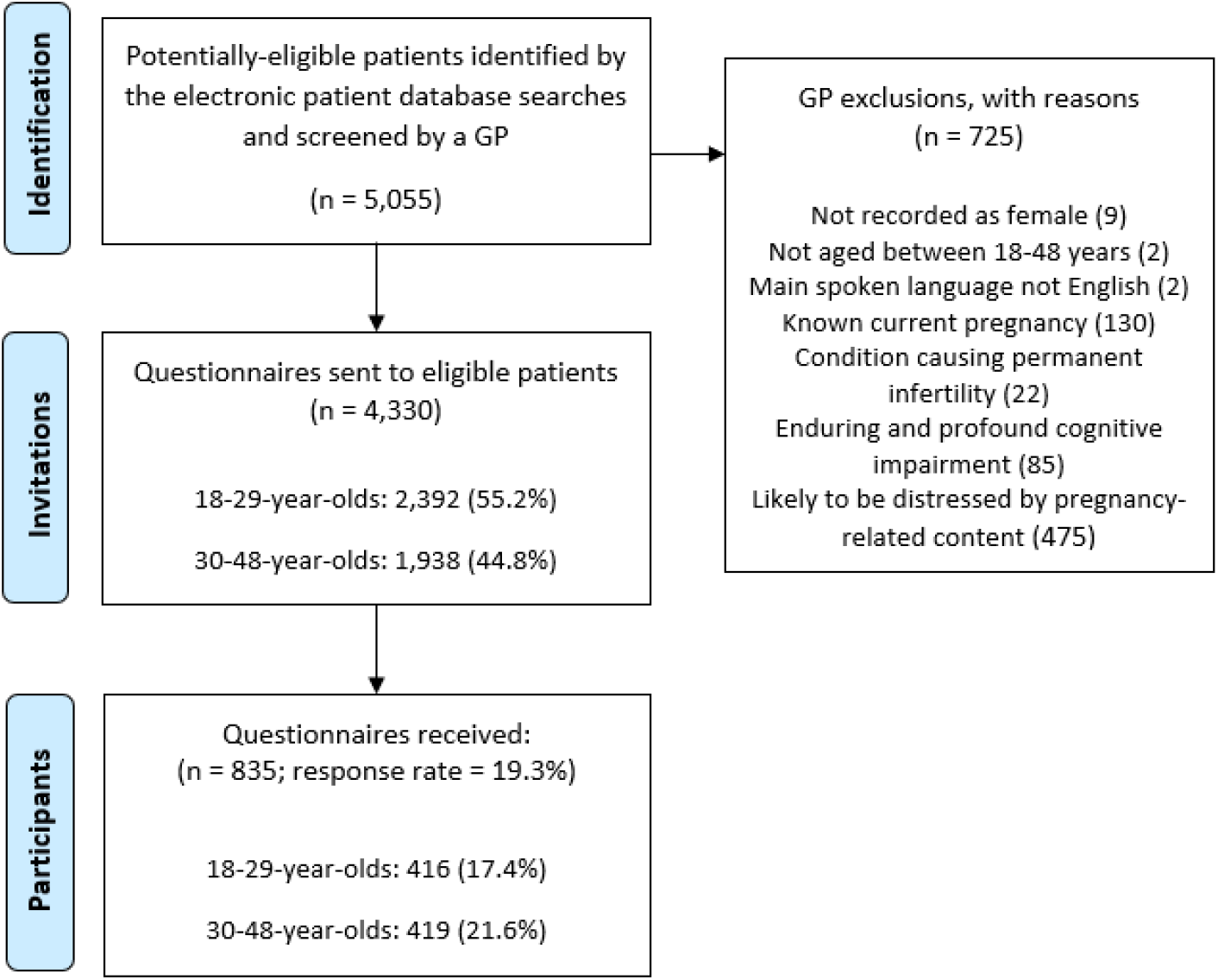
Study enrolment flowchart

### Descriptive data

Table 1 shows there was an even split of 18-29 and 30-48-year-old participants. A third (32.1%) of the study population had household incomes below £32,000, compared with 43% of the UK population (37). A greater proportion of the study sample were UK-born (91.9 vs 84.3%), identified as White (92.6 vs 84.8%), and were university graduates (68.6 vs 42%) than the national population. Approximately half (48.4%) of participants had previously been pregnant, and a third (38.8%) of participants reported at least one live birth. All five participants who reported a stillbirth also reported a live birth(s).

**Table 1:**
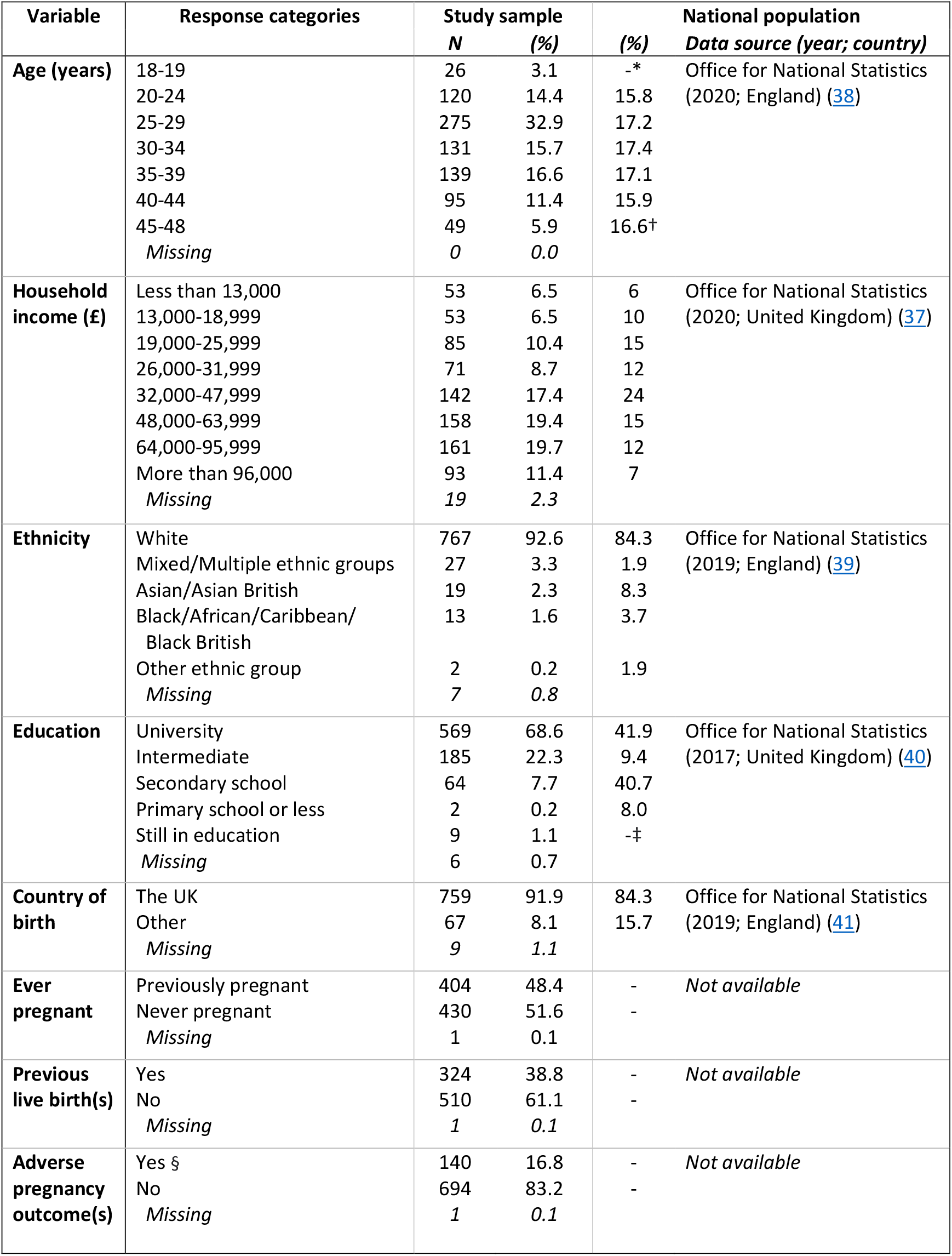
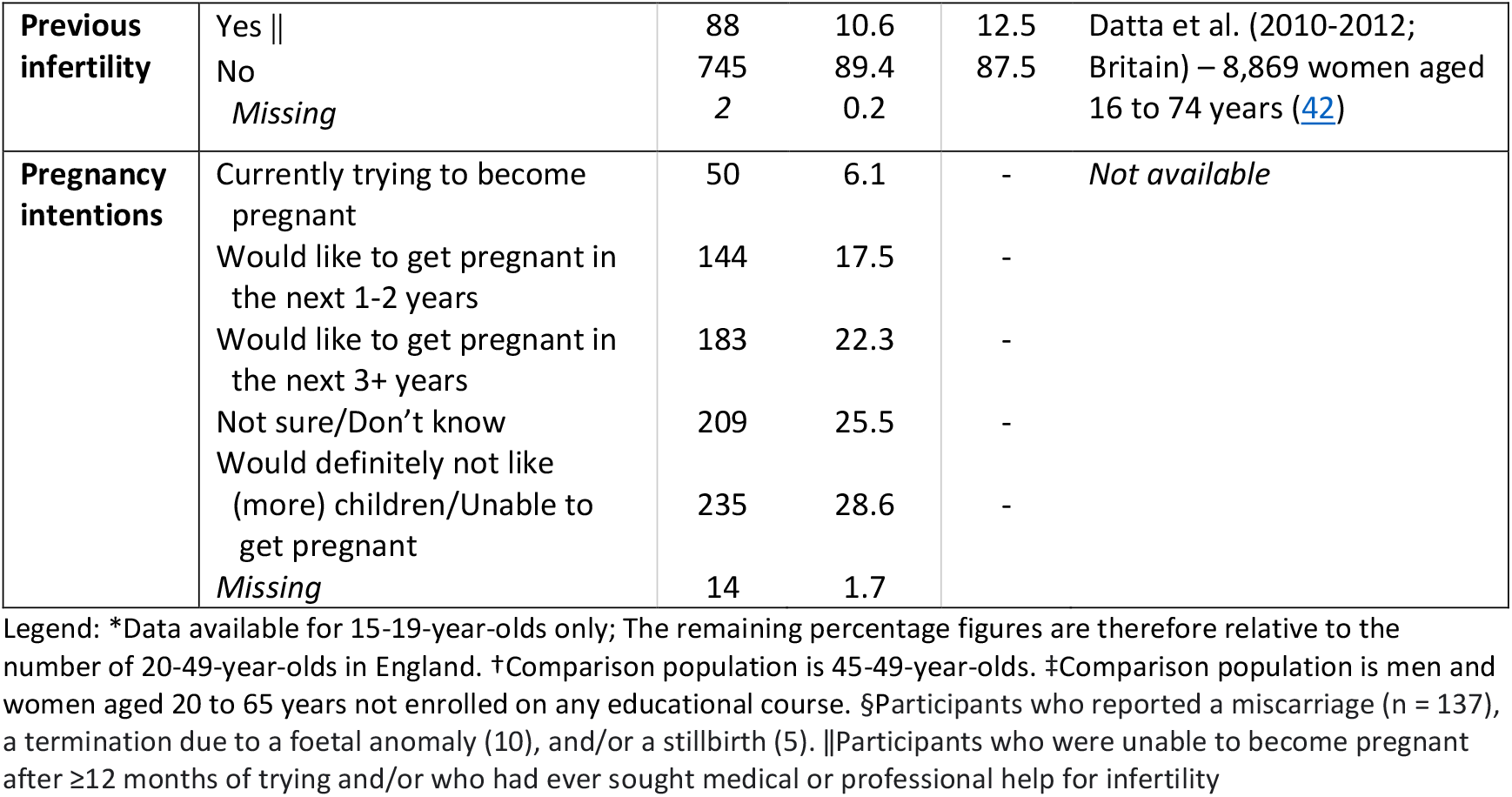
Characteristics of the study population, with comparison to the national population.

### Knowledge

Table 2 shows maternal diet and physical activity were the only two risk factors listed by the majority of participants (86.0% and 79.2%, respectively). Less than half of women listed maternal BMI/weight, folic acid, abuse, age, and interpregnancy intervals. No participants mentioned interpregnancy weight change or avoiding excess iron intake. The preconception exposures most commonly listed for which there was *no* high, moderate or low certainty evidence of an association(s) with an adverse pregnancy, birth or postpartum outcome(s) in Daly et al. (5) were: alcohol consumption (89.7%), smoking (89.3%), stress (51.6%), substance abuse (48.3%), vitamins (without explicit mention of folic acid; 34.9%), mental health/self-care (33.1%) and social support and relationships (25.3%). The full list is presented in Additional file 4.

**Table 2:**
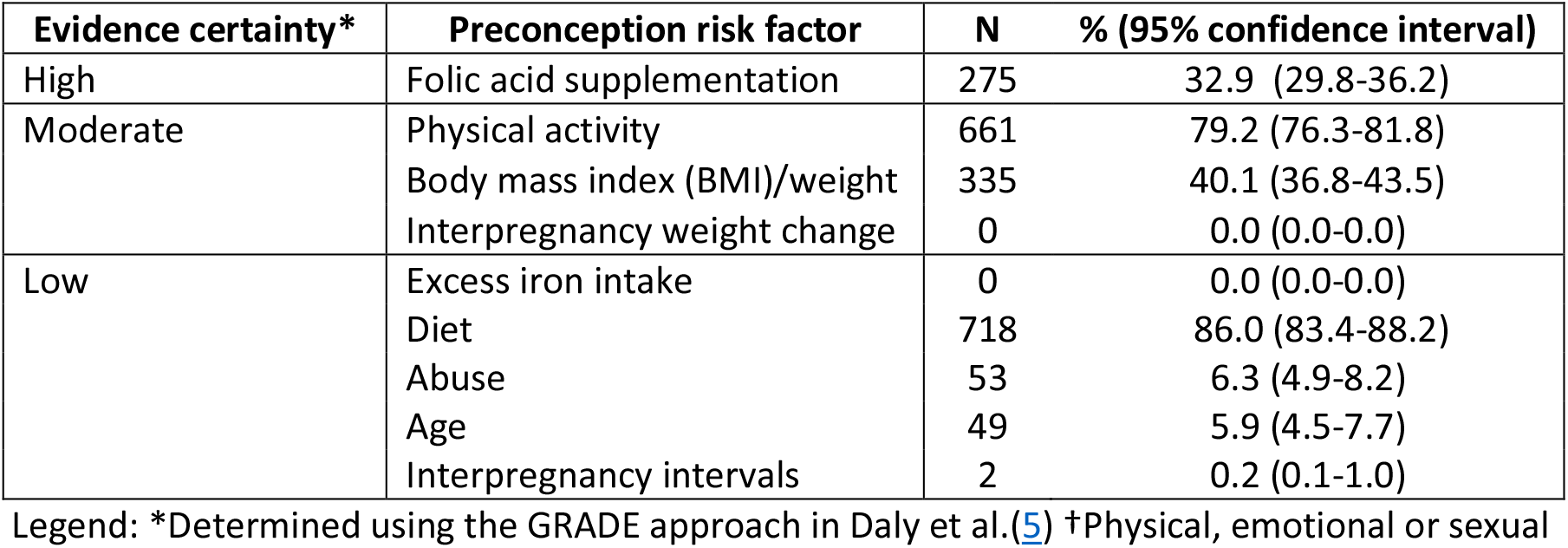
Participant knowledge levels of preconception risk factors.

Table 3 shows the adjusted associations between participant characteristics and knowledge of preconception risk factors. Interpregnancy weight change, excess iron intake and interpregnancy intervals were excluded from these analyses as zero and two participants listed these factors, respectively. The proportions of participants who listed each risk factor, by participant characteristic, are presented in Additional file 5. After adjustment, older age, university education, pregnancy desire, gravidity, prior live birth(s) and adverse pregnancy outcome(s) were associated with knowledge of the benefit of preconception folic acid supplementation. The likelihood of listing BMI as a risk factor rose with increasing household income.

**Table 3:**
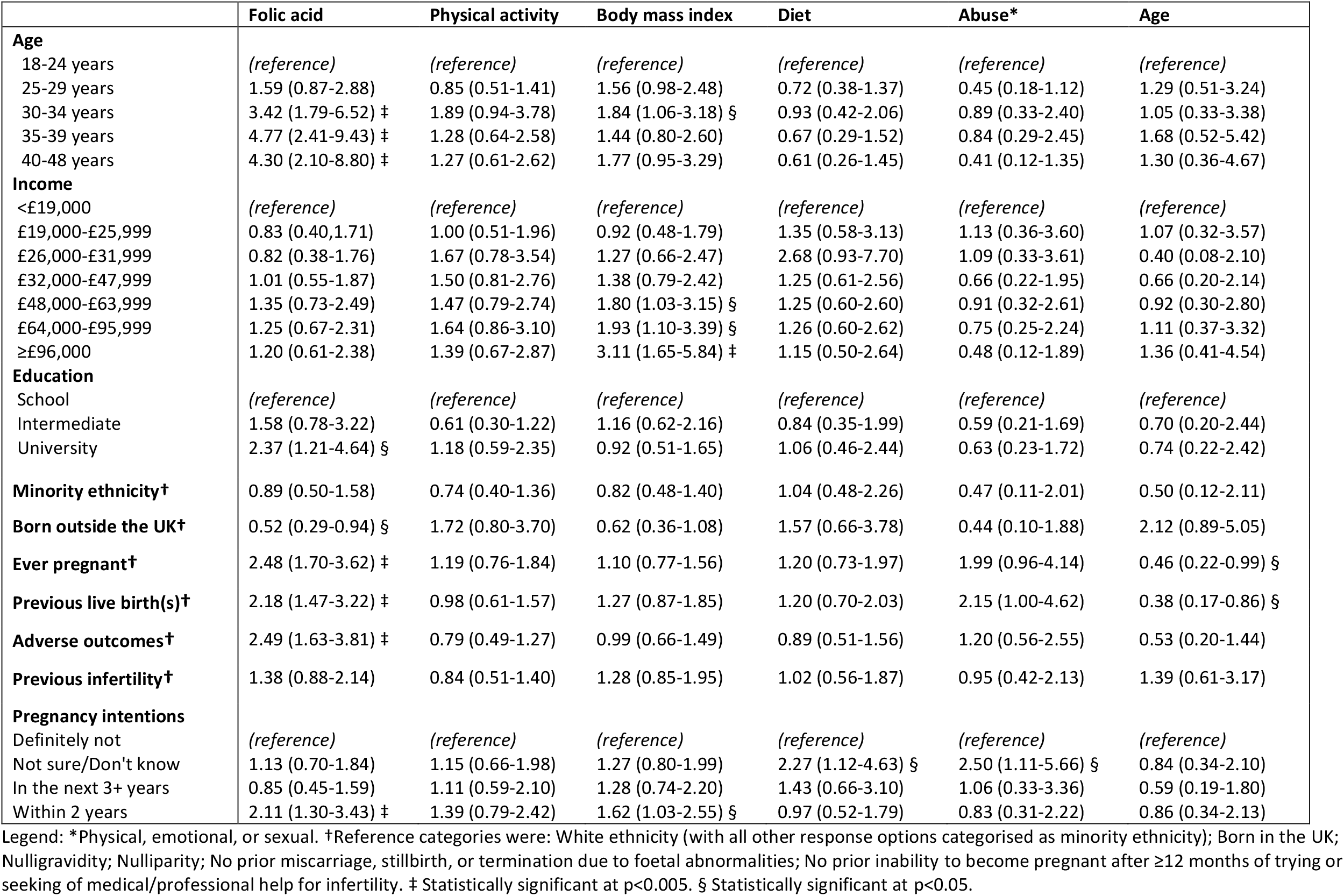
Adjusted odds ratios for associations between participant characteristics and knowledge of preconception risk factors.

### Attitudes

Over a third of participants felt they were slightly or not at all aware of preconception health risk factors, and almost half of participants were either slightly or not at all interested in knowing more about preconception health (Additional file 6). Few participants disagreed that maternal preconception health can affect maternal and infant health outcomes or that they could make positive preconception changes, or reported that they would be unlikely to make these changes. Table 4 shows that after adjustment, women over 30 years, gravid and parous women, and women who reported adverse pregnancy outcomes felt more aware of preconception risk factors. Higher-income and minority ethnic women rated the importance of preconception health higher. Women under 40 years, minority ethnic, gravid and parous women, and women wishing to become pregnant were more interested in preconception health education. Women aged 40 and over had greater belief they could make positive preconception changes.

**Table 4:**
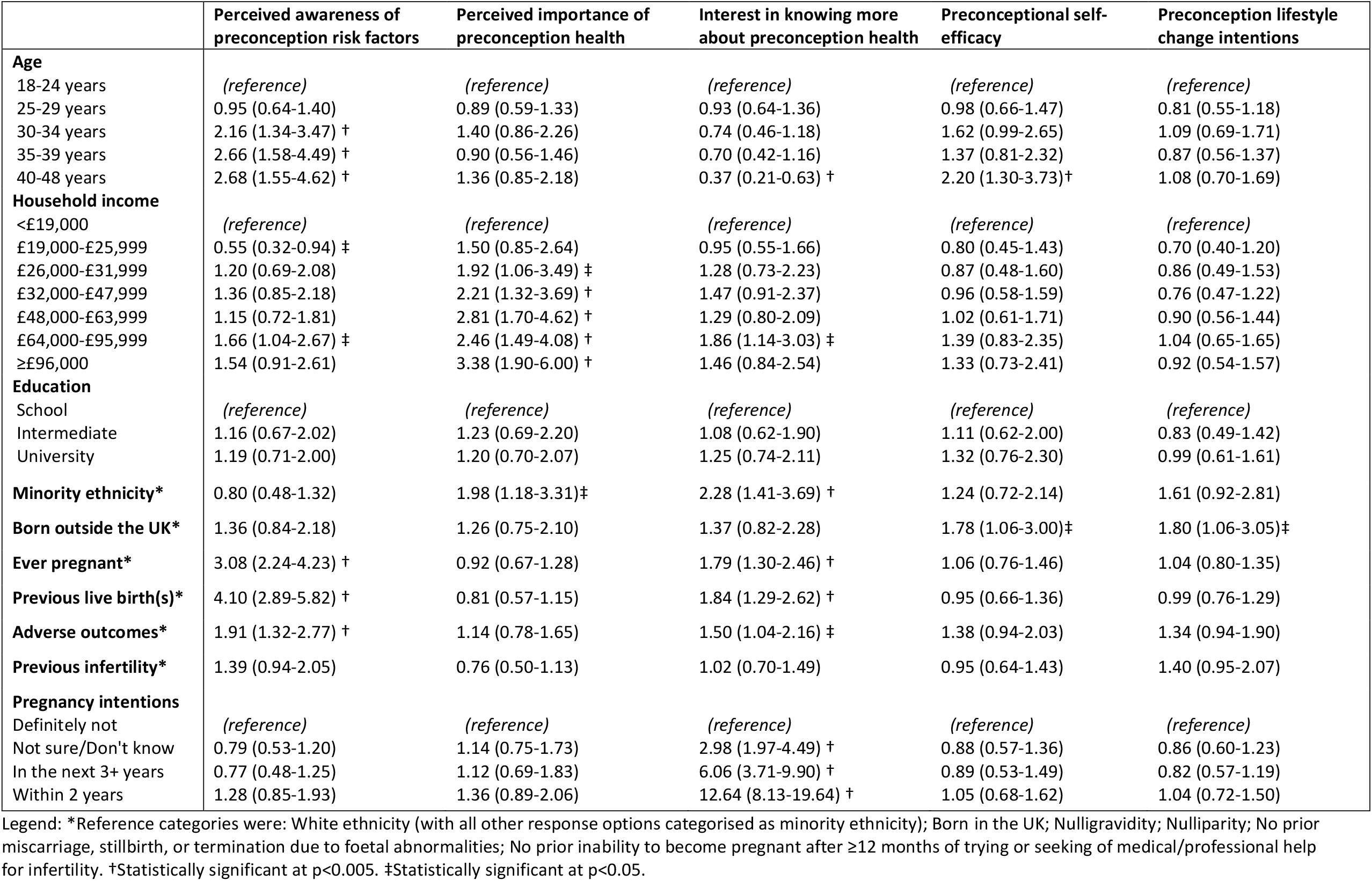
Adjusted odds ratios for associations between participant characteristics and attitudes towards preconception health.

|  | Perceived awareness of preconception risk factors | Perceived importance of preconception health | Interest in knowing more about preconception health | Preconceptional self-efficacy | Preconception lifestyle change intentions |
| --- | --- | --- | --- | --- | --- |
| **Age** |  |  |  |  |  |
| 18-24 years | (reference) | (reference) | (reference) | (reference) | (reference) |
| 25-29 years | 0.95 (0.64-1.40) | 0.89 (0.59-1.33) | 0.93 (0.64-1.36) | 0.98 (0.66-1.47) | 0.81 (0.55-1.18) |
| 30-34 years | 2.16 (1.34-3.47) † | 1.40 (0.86-2.26) | 0.74 (0.46-1.18) | 1.62 (0.99-2.65) | 1.09 (0.69-1.71) |
| 35-39 years | 2.66 (1.58-4.49) † | 0.90 (0.56-1.46) | 0.70 (0.42-1.16) | 1.37 (0.81-2.32) | 0.87 (0.56-1.37) |
| 40-48 years | 2.68 (1.55-4.62) † | 1.36 (0.85-2.18) | 0.37 (0.21-0.63) † | 2.20 (1.30-3.73)† | 1.08 (0.70-1.69) |
| **Household income** |  |  |  |  |  |
| <£19,000 | (reference) | (reference) | (reference) | (reference) | (reference) |
| £19,000-£25,999 | 0.55 (0.32-0.94) ‡ | 1.50 (0.85-2.64) | 0.95 (0.55-1.66) | 0.80 (0.45-1.43) | 0.70 (0.40-1.20) |
| £26,000-£31,999 | 1.20 (0.69-2.08) | 1.92 (1.06-3.49) ‡ | 1.28 (0.73-2.23) | 0.87 (0.48-1.60) | 0.86 (0.49-1.53) |
| £32,000-£47,999 | 1.36 (0.85-2.18) | 2.21 (1.32-3.69) † | 1.47 (0.91-2.37) | 0.96 (0.58-1.59) | 0.76 (0.47-1.22) |
| £48,000-£63,999 | 1.15 (0.72-1.81) | 2.81 (1.70-4.62) † | 1.29 (0.80-2.09) | 1.02 (0.61-1.71) | 0.90 (0.56-1.44) |
| £64,000-£95,999 | 1.66 (1.04-2.67) ‡ | 2.46 (1.49-4.08) † | 1.86 (1.14-3.03) ‡ | 1.39 (0.83-2.35) | 1.04 (0.65-1.65) |
| ≥£96,000 | 1.54 (0.91-2.61) | 3.38 (1.90-6.00) † | 1.46 (0.84-2.54) | 1.33 (0.73-2.41) | 0.92 (0.54-1.57) |
| **Education** |  |  |  |  |  |
| School | (reference) | (reference) | (reference) | (reference) | (reference) |
| Intermediate | 1.16 (0.67-2.02) | 1.23 (0.69-2.20) | 1.08 (0.62-1.90) | 1.11 (0.62-2.00) | 0.83 (0.49-1.42) |
| University | 1.19 (0.71-2.00) | 1.20 (0.70-2.07) | 1.25 (0.74-2.11) | 1.32 (0.76-2.30) | 0.99 (0.61-1.61) |
| **Minority ethnicity\*** | 0.80 (0.48-1.32) | 1.98 (1.18-3.31)‡ | 2.28 (1.41-3.69) † | 1.24 (0.72-2.14) | 1.61 (0.92-2.81) |
| **Born outside the UK\*** | 1.36 (0.84-2.18) | 1.26 (0.75-2.10) | 1.37 (0.82-2.28) | 1.78 (1.06-3.00)‡ | 1.80 (1.06-3.05)‡ |
| **Ever pregnant\*** | 3.08 (2.24-4.23) † | 0.92 (0.67-1.28) | 1.79 (1.30-2.46) † | 1.06 (0.76-1.46) | 1.04 (0.80-1.35) |
| **Previous live birth(s)\*** | 4.10 (2.89-5.82) † | 0.81 (0.57-1.15) | 1.84 (1.29-2.62) † | 0.95 (0.66-1.36) | 0.99 (0.76-1.29) |
| **Adverse outcomes\*** | 1.91 (1.32-2.77) † | 1.14 (0.78-1.65) | 1.50 (1.04-2.16) ‡ | 1.38 (0.94-2.03) | 1.34 (0.94-1.90) |
| **Previous infertility\*** | 1.39 (0.94-2.05) | 0.76 (0.50-1.13) | 1.02 (0.70-1.49) | 0.95 (0.64-1.43) | 1.40 (0.95-2.07) |
| **Pregnancy intentions** |  |  |  |  |  |
| Definitely not | (reference) | (reference) | (reference) | (reference) | (reference) |
| Not sure/Don't know | 0.79 (0.53-1.20) | 1.14 (0.75-1.73) | 2.98 (1.97-4.49) † | 0.88 (0.57-1.36) | 0.86 (0.60-1.23) |
| In the next 3+ years | 0.77 (0.48-1.25) | 1.12 (0.69-1.83) | 6.06 (3.71-9.90) † | 0.89 (0.53-1.49) | 0.82 (0.57-1.19) |
| Within 2 years | 1.28 (0.85-1.93) | 1.36 (0.89-2.06) | 12.64 (8.13-19.64) † | 1.05 (0.68-1.62) | 1.04 (0.72-1.50) |

Legend: \*Reference categories were: White ethnicity (with all other response options categorised as minority ethnicity); Born in the UK; Nulligravidity; Nulliparity; No prior miscarriage, stillbirth, or termination due to foetal abnormalities; No prior inability to become pregnant after ≥12 months of trying or seeking of medical/professional help for infertility. †Statistically significant at p<0.005. ‡Statistically significant at p<0.05.

### Acceptability and use of intervention delivery channels

Figure 2 shows the most acceptable places and settings for preconception health information provision were: preconception health websites/apps (99.5% ‘acceptable’), printed material in healthcare settings (98.6%), health education in schools (94.4%), television (91.9%), pregnancy tests (91.0%), and social media (88.2%). Figure 3(a) shows that, of the delivery channels involving people, family/partners (96.3%), doctors, midwives and nurses (86.8-97.0%), friends (86.5%), and sexual health/family planning staff (79.7%) were the most acceptable. Dentists (23.9%) and hairdressers/ beauticians (18.1%) were the least acceptable. Figure 3(b) shows that many participants reported ‘never’ having contact with: community/family support workers (87.0%), health visitors (57.4%), midwives (55.6%), sexual health/family planning staff (49.2%), and obstetricians/gynaecologists (48.5%). For all remaining options, at least 85% of participants reported contact in the last 3 years.

**Figure 2:**
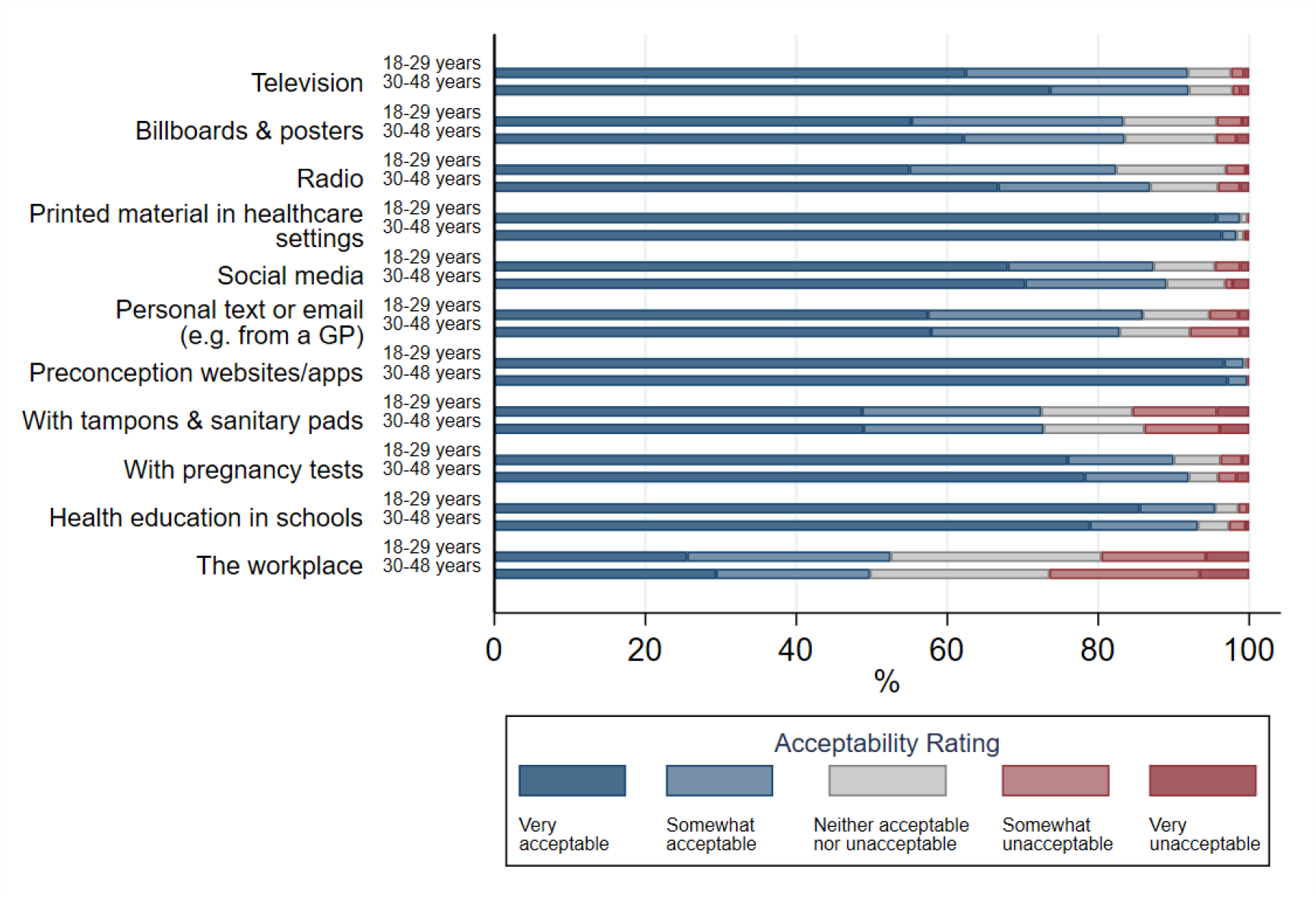
Acceptability of providing preconception health information in various places and settings

**Figure 3:**
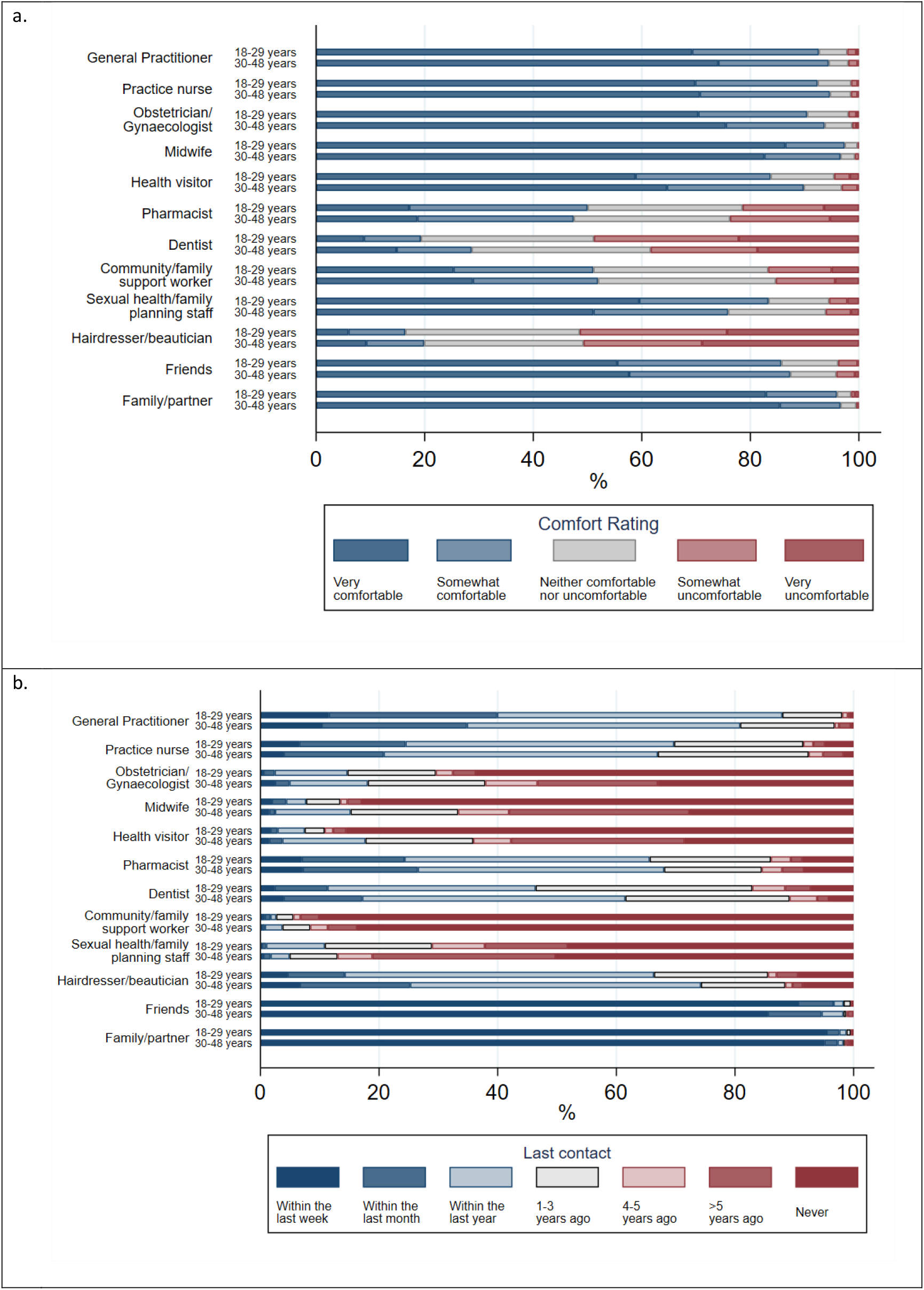
Acceptability of discussing preconception health (a) and last contact (b) with various people

## Discussion

Preconception health is an increasing priority for policy makers, public health and clinicians. This is the first UK study since 1998 to investigate women’s knowledge of and attitudes towards preconception health, and the first to explore the acceptability of both healthcare- and community-based intervention delivery methods. Preconception health knowledge was generally low. Most women were unaware that a lack of folic acid supplementation, BMI, interpregnancy weight change, abuse, advanced age, excess iron intake and interpregnancy intervals are maternal preconception risk factors. We identified that younger women and nulligravid women were less aware of the benefit of preconception folic acid supplementation, and women with lower household incomes were less aware of the importance of preconception BMI. Women’s attitudes towards preconception health were generally positive, though almost half reported low interest in preconception health education. Younger women were more interested in this, whereas nulligravid women were less interested. Women with lower household incomes rated the importance of preconception health lower. Most women reported recent contact with GPs, practice nurses, pharmacists, and family/partners, and considered these acceptable intervention delivery channels. Websites/apps, printed healthcare materials, schools, pregnancy tests, television and social media were also considered acceptable settings for preconception health information provision.

### Limitations

Our study has some limitations relating to its sample, response rate, and administration. As responses were self-reported, some women may have consulted online material to inform their questionnaire responses. This may have resulted in an overestimation of knowledge, as women might have listed preconception risk factors they would not have otherwise been aware of. Administration by a researcher may have led to underreporting, however, due to the sensitive nature of some of the questions. Regarding the study’s sample, the overrepresentation of university graduates and White, UK-born and high-income women limits the generalisability of the study’s findings to the UK population. The low number of responses from minority ethnic women and women born outside the UK also limited our ability to detect precise outcome estimates for these groups. Moreover, if preconception health was of greater relevance to the minority of women who took part than those who did not, this may have introduced a self-selection bias, where knowledge was greater and attitudes more favourable. This is less likely to have affected the reported associations, however (43).

A further potential limitation is our choice of preconception risk factors. While we felt it was prudent to select preconception risk factors based on the certainty of the evidence for their associations with adverse pregnancy, birth and postpartum outcomes, there are arguably other valid reasons for practicing or avoiding other exposures preconceptionally. These include the avoidance of antenatal exposure arising from late pregnancy recognition, or the difficulty involved in making immediate changes to established habits in pregnancy (15). From this perspective, exposures such as smoking and alcohol consumption, listed by most participants, could be considered appropriate responses. So too could responses such as mental health/self-care and social support and relationships, as these may be important determinants and facilitators of positive preconception behaviours. We have therefore included the full list of participant-suggested risk factors in Additional file 4.

### Interpretation

The findings of this study do not replicate those from the last assessment of preconception knowledge and attitudes in the United Kingdom, performed 25 years ago (20). In that study, reproductive-aged women in England were “generally well informed” about preconception health. We found low knowledge of preconception risk factors in the present study, particularly of preconception-specific factors like folic acid supplementation and interpregnancy intervals. They found 40% of women considered preconception care to be ‘essential’. In our study we found 92.7% of our participants perceived preconception health to be important. Birth outside the UK, minority ethnicity, lack of higher education and nulligravidity were the strongest correlates of low preconception health knowledge in Wallace and Hurwitz (20), but we found only lack of higher education and nulligravidity were associated with this lack of knowledge, along with younger age and lower household incomes. These differences may be due to the lack of multivariate statistical methods in Wallace and Hurwitz (20), or our study having fewer women from ethnic minority groups but recruiting more general practices from socioeconomically deprived areas.

Our findings also contrast with those of Stern et al. (19), the only other study to have assessed preconception health knowledge without naming risk factors in their questionnaire. A minority of participants in the present study listed folic acid, though fewer respondents in Stern et al. listed this risk factor (33% vs 4%). This may reflect actual differences in knowledge between Swedish and English women, or be attributable to Stern et al.’s sampling of university students only, as younger age was associated with a lack of knowledge of this risk factor in both studies. Both studies also found lower educational attainment and lack of pregnancy desire to correlate with low knowledge of preconception health. Unlike the present study, however, gravidity was not associated with knowledge in Stern et al., though this may be due to the low number of gravid women (17) in their control groups. Moreover, fewer women in the present study reported that they were unlikely to make preconception lifestyle changes than in Stern et al. (6.8% vs 14%). This may reflect either national attitudinal differences or differential lifestyle behaviours amongst students.

### Future research

Future research should seek to identify explanations for why some women are less interested and less aware of preconception health, perceive it to be less important, and are less likely and less confident in their ability to make preconception health changes. Research has implicated factors like cost and time availability as barriers to other health behaviours, like healthy eating and physical activity (44), but it is yet unclear whether these apply to preconception health specifically. Moreover, evidence from systematic reviews suggests interventions targeting women’s knowledge of and attitudes towards preconception health can be effective in improving knowledge and health behaviours (13, 15), and there is some evidence from services and interventions that effecting positive changes in these factors can relate to improved maternal health and pregnancy outcomes (45-47). We identify here acceptable delivery methods for interventions targeting these factors; Future research should explore conditions that might further affect the acceptability of these methods, as this may increase the likelihood of intervention effectiveness (48) and stakeholder buy-in (49).

Future research should also explore the acceptability of low-agency population interventions, such as food fortification and activity-promoting environments, that may work in tandem with interventions seeking to provide advice, guidance, and encouragement around preconception health (50).

## Conclusions

Our findings highlight the need for public health interventions promoting the importance of preconception health and knowledge of lesser-known preconception risk factors such as folic acid supplementation, maternal BMI, and interpregnancy intervals, particularly amongst younger and nulliparous women, and women with lower incomes. Younger women were more interested in learning more about preconception health and may therefore be more receptive to these interventions. Healthcare providers and settings were widely viewed as acceptable intervention delivery methods, but we also found that methods in community settings, such as family/partners, social media, websites and apps, health education in schools and pregnancy tests, were acceptable. There is arguably a need for greater consideration to these methods, given that many women – younger women in particular - reported never having contact with most healthcare provider groups. Moreover, healthcare-based methods are less likely to reach individuals at risk of an unplanned pregnancy, or whose pregnancy intentions are not known to their healthcare provider (18), and women are more likely to receive information on preconception health from non-healthcare sources (14). This suggests that methods such as population-wide information campaigns using social media and health education in schools may have greater reach in accessing younger and nulliparous women.

## Supporting information

STROBE checklist

## Data Availability

All data produced in the present study are available upon reasonable request to the authors

## List of abbreviations

*AHEI:*: Alternate Healthy Eating Index
*BMI:*: Body Mass Index
*GP:*: General Practitioner
*Natsal-3:*: National survey of sexual attitudes and lifestyles

## Declarations

### i. Ethics approval and consent to participate

The study received ethical approval from the South West – Frenchay Research Ethics Committee (committee reference number 19/SW/0235) prior to its conduct. All methods were carried out in accordance with the Declaration of Helsinki and all other relevant regulations and guidelines. The first page of the questionnaire informed participants that to take part in the study, they were required to read and tick each of four statements indicating that they: (i) had read the relevant version of the Participant Information Sheet and had the opportunity to ask questions and have these answered fully; (ii) understood that their participation was voluntary; (iii) understood that the data they provided could be stored and used in its anonymised form for reports, publications, and/or teaching materials from the research and by other researchers for other research; (iv) agreed to take part in the study. Informed consent was obtained from all participants. Contact details for the study team were included in the study invitation letter to give patients the opportunity to ask questions before participating. Personally identifiable data will not be shared outside the study team.

### ii. Consent for publication

Not applicable.

### v. Authors’ contributions

RK conceived the idea of the study and all authors were involved in its design. MD was responsible for: preparing the study’s ethical review application; recruiting and liaising with participating GP practices; entering and preparing the study data; performing the statistical analyses; and writing the final report, with guidance and input from RK, JW and JS. All authors read, edited and approved the final manuscript.

## vi. Acknowledgements

The authors thank the women who took part in this study, the staff of the general practices who acted as the study’s participant identification centres, and NIHR CRN West of England who helped us to recruit these practices and funded the study’s service support costs. We also wish to thank: Rona Campbell, Deborah Lawlor, Gemma Morgan and Rhiannon Macefield, who provided guidance on the study’s design; Mike Bell at NIHR ARC West, who facilitated the study’s public involvement; and Judith Stephenson, Chandni Maria Jacob, Deirdre de Barra and Jo Williams, who reviewed the study’s protocol and materials prior to its conduct.

