## Supplemental material for "Women’s knowledge, attitudes and views of preconception health and intervention delivery methods: A cross-sectional survey"

**Additional file 1: Questions stems, response options, sources, psychometric properties, and modifications made to the survey’s questionnaire items**

i. ***Exposure variables***

| **Variable** | **Question stem and response options** | **Item source, existing psychometric properties, and modifications made** |
| --- | --- | --- |
| Age | *What is your age?*  18-19, 20-24, 25-29, 30-34, 35-39, 40-44, or 45-48 years | Determined using the Government Statistical Service’s Age Band Group 1 D harmonised principle groupings (1), with the final option modified in line with the survey’s upper age limit. |
| Educational attainment | *What is the highest level of education you have completed?*  University, Intermediate between secondary school and university (e.g. technical training), Secondary school, Primary school (or less), Still in education /not yet finished | Determined using an adaption of the ‘*what is the highest level of education you have completed?*’ item used by the World Health Organisation’s MONICA project (2), with the addition of the response option ‘*still in education/not yet finished’*. No validity or reliability properties were reported for the questionnaire used by the MONICA project (2). |
| Ethnicity | *What is your ethnic group? Circle ONE option that best describes your ethnic group or background*  White, Mixed/Multiple ethnic groups, Asian/Asian British, Black/African/Caribbean/Black British, Other ethnic group, please describe | Determined using the Government Statistical Service’s recommended ethnic group question for use in surveys in England (1). Only the higher-level ethnicity groupings were used as response options, but with ‘*other ethnic group*’ replaced with ‘*other ethnic group, please describe:..*’ to allow participants who were unsure of their ethnicity grouping to be categorised by the study team. This question arose from a two-year cross-government stakeholder consultation programme (3). |
| Country of birth | *What is your country of birth?*  The UK, Other (please write) | Determined using the Government Statistical Service’s harmonised principle items for ‘*country of birth*’ and ‘*year and of arrival in the UK*’ (1). |
| Household income | *What was your total household income last year? (before taxes or deductions)*  Less than £13,000, £13,000-£18,999, £19,000-£25,999, £26,000-£31,999, £32,000-£47,999, £48,000-£63,999, £64,000-£95,999, More than £96,000 | Determined using a self-developed item (‘*what was your total household income last year? (before taxes or deductions)*’), with response options derived from the Office of National Statistics’ ‘*Gross household income, UK, financial year ending 2018*' report (4), to allow a comparison between the survey respondents and the wider UK population. |

| **Variable** | | **Question stem and response options** | **Item source, existing psychometric properties, and modifications made** | |
| --- | --- | --- | --- | --- |
| Gravidity | | *Have you ever been pregnant before? (circle ONE option)*  Yes, I am pregnant now and this is my first pregnancy, No | Determined using an adaption of the ‘*Everpreg*’ item in the third British national survey of sexual attitudes and lifestyle (Natsal-3) questionnaire (5). The response option ‘*I am pregnant now and this is my first pregnancy*’ was added to identify participants who had experience of becoming pregnant but would not be expected to report pregnancy outcomes for the proceeding item. Participants who selected ‘*no*’ were classed as nulligravida. The Natsal-3 questionnaire has been validated for use in UK populations through cognitive interviewing and two large pilot studies (5). | |
| Previous Live Birth(s)  Adverse Pregnancy Outcomes | | *How many of each of the following have you had? (enter ‘0’ for anything you have not experienced)*  Miscarriage (or an ectopic pregnancy), Termination or abortion due to foetal abnormalities, Termination or abortion for other reasons, Stillbirth (a baby born after 24 weeks of pregnancy that died before birth), A live birth/baby | Determined using an adaption of the Natsal-3 questionnaire’s ‘PregOL’ item (5). ‘*What was the outcome of that pregnancy?*’ was replaced with ‘*How many of each of the following have you had? (enter ‘0’ for anything you have not experienced)*’ as the question stem. The ‘*live birth (one child)*’ and ‘*live birth (more than one child)*’ response options were collapsed to form a singular ‘*live birth(s)*’ option, and the ‘*termination or an abortion*’ response option was split into separate ‘…*due to foetal abnormalities*’ and ‘…*for other reason(s)*’ options. Participants who reported one or more miscarriages, stillbirths and/or terminations due to foetal abnormalities were categorised as having experienced an adverse pregnancy outcome. | |
| Previous infertility | | *Circle ONE number for each of the following two questions: Have you ever had a time, lasting 12 months or longer, when you and a partner were trying for a pregnancy but it didn't happen? Have you or a partner ever sought medical or professional help about infertility?*  Yes, No | Determined using the Natsal-3 questionnaire’s ‘*Infrt1y*’ and ‘*MedHelp*’ items (5). Participants who respond ‘*yes*’ to either of these items were categorised as having experienced fertility issues | |
| **Variable** | **Question stem and response options** | | | **Item source, existing psychometric properties, and modifications made** |
| Pregnancy intentions | *Which of these statements best describes the way you feel about having (more) children? (Circle ONE number)*  I would definitely like (more) children, and I’m currently trying,  I would definitely like (more) children, I’m not currently trying, but I would like to get pregnant in the next 1-2 years  I would definitely like (more) children. I’m not currently trying, but I would like to get pregnant in the next 3+years,  I might like (more) children in the future, I’m not sure yet I would definitely not like (more) children,  Don’t know, I’m not able to get pregnant | | | Determined using an adaption of Natsal-3’s ‘*FertInt*’ item (5). The response option ‘*I would definitely like (more) children, but I’m not currently trying*’ was split to form separate ‘*I would definitely like (more) children. I’m not currently trying, but I would like to get pregnant in the next 1-2 years*’ and ‘…*in the next 3+ years*’ options, to better capture the immediacy of participants’ pregnancy desire. The option ‘*I’m not able to get pregnant*’ was also added following validity testing. |

ii. ***Outcome variables***

| **Variable** | **Question stem and response options** | **Item source, existing psychometric properties, and modifications made** |
| --- | --- | --- |
| Knowledge of preconception risk factors | *List up to 5 things you think a woman can DO (or START or CONTINUE doing) BEFORE pregnancy to help her to have a healthy pregnancy and healthy baby*  *List up to 5 things you think a woman can STOP doing (or AVOID) BEFORE pregnancy to help her to have a healthy pregnancy and healthy baby*  *List ANY OTHER things BEFORE pregnancy that might affect whether a woman has a healthy pregnancy and baby. These might be about her health, her life or her circumstances before pregnancy, or other things.* | Only one identified study – Stern et al. (6) - assessed knowledge of preconception risk factors using an open-ended, free-text question. The authors asked only about individual-level lifestyle changes and assigned ‘points’ for eight risk factors. The remaining studies asked specific questions about specific preconception exposures (e.g. “*which of the following actions have you heard are the most important things for women to do before they get pregnant?... Avoid smoking cigarettes”* (7)), bestowing salience to these factors and likely resulting in an overestimation of knowledge by eliciting desired responses.  The item from Stern et al. (6), piloted for use in a population of reproductive-age Swedish women and reviewed by academics, clinicians and laypeople, was therefore modified to form multiple items asking participants to list things that, before pregnancy, can be: (1) done, started or continued; (2) stopped or avoided; or that (3) relate to a woman’s life, circumstances or health, which might affect pregnancy outcomes.  For each exposure identified in a recent umbrella review of preconception exposures (8), for which high, moderate or low certainty evidence of a preconceptional association with an adverse pregnancy, birth and postpartum outcome(s) was found, participants were scored as ‘yes’ or ‘no’ depending on whether they had listed it. |
| Perceived awareness of preconception health behaviours | *How aware are you of the positive behaviours and other actions women can take BEFORE pregnancy to help to have a healthy pregnancy and a healthy baby? (Circle ONE of the options)*  Very aware, Moderately aware, Slightly aware, Not aware at all | Of the survey studies identified, the item ‘*to what extent do you feel you are aware of the positive behaviours and other actions pregnant women can take to increase their odds of having a healthy pregnancy and a healthy baby?*’ in Delgado et al. (9), scored on a four-point Likert scale, was deemed the most suitable for this variable. The beginning of the question stem was modified to ‘*How aware are you of…can take BEFORE pregnancy to help to have a…*’ to make the wording less complex, less leading, and more specific to the preconception period.  No validity or reliability properties were reported for the questionnaire used in Delgado et al. (9) |

| **Variable** | **Question stem and response options** | **Item source, existing psychometric properties, and modifications made** |
| --- | --- | --- |
| Perceived importance of preconception health | *Please rate how much you agree with the following (circle ONE option): “A woman’s health BEFORE pregnancy can affect the health of her and her baby during and after pregnancy”*  Strongly agree, Agree, Neither agree nor disagree, Disagree, Strongly disagree | Of the survey studies identified, the item ‘*A woman’s health before conception can have serious consequences to the health of the baby*’ in Best Start Resource Center’s questionnaire (10) was deemed the most suitable for this variable. Its question stem was modified to ‘*A woman’s health BEFORE pregnancy can affect the health of her and her baby during and after pregnancy*’ to be more lay and applicable to both child and maternal pregnancy outcomes.  No validity or reliability properties were reported for the questionnaire used by Best Start Resource Center (10). |
| Interest in preconception health education | *How interested are you in knowing more about pre-pregnancy health? (Circle ONE option)*  Very interested, Moderately interested, Slightly interested, Not at all interested | Of the survey studies identified, the item ‘*Are you interested in receiving preconception health education?*’ in Frey et al. (11) was deemed the most suitable for this variable. Its question stem was modified to ‘*How interested are you in knowing more about pre-pregnancy health?*’ to be less leading and complex, and the response options were changed to ‘*Very*’, ‘*Moderately*’, ‘*Slightly*’ and ‘*Not at all*’ interested, in line with the response options for ‘*Perceived Importance of Preconception Health’*. |
| Preconcept-ional self-efficacy | *Imagine that you were planning to become pregnant within 6 months. How much do you agree with the following statement? (circle ONE option): “There are things I can do before I become pregnant to help make sure my child is born healthy”*  Strongly agree, Agree, Neither agree nor disagree, Disagree, Strongly disagree | Determined using the Preconceptional Control item in Weisman et al. (12). The item introduction (‘*Imagine that you were planning to become pregnant within 6 months*’) was derived from the ‘*Pregnancy planning as cue to action*’ item in Stern et al. (6). Scores on this item have been positively associated with folic acid use and receipt of pregnancy planning counselling (12). Dose-response effects were also reported for this item, for each additional session attended of a behaviour change intervention to improve the preconceptional and inter-conceptional health of women (odds ratio [OR] 1.31, *p*=0.002), alongside significant dose-response effects for several health behaviours such as checking food labels for nutritional information (OR 1.16, *p*=0.015), partaking in relaxation exercise or meditation for stress management (OR 1.24, *p*= 0.009), and daily use of a multivitamin with folic acid (OR 1.45, *p*=0.009) (13). |

| **Variable** | **Question stem and response options** | **Item source, existing psychometric properties, and modifications made** |
| --- | --- | --- |
| Preconception health intentions | *Imagine that you were planning to become pregnant within 6 months. How likely is it that you would make any lifestyle changes during these 6 months, in preparation for pregnancy? (circle ONE)*  Very likely, Quite likely, Neither likely nor unlikely, Quite unlikely, Very unlikely | Determined using the ‘*Pregnancy planning as cue to action’* item in Stern et al. (6), scored on a five-point Likert scale.  The questionnaire used in Stern et al. (6) was piloted for use in a population of reproductive-age Swedish women |
| Intervention delivery method acceptability  (People/staff groups) | *Please rate how comfortable you would be with discussing health before pregnancy and your personal pregnancy plans with each of the following people (circle ONE of the following options for each person)*  Very comfortable, Somewhat comfortable, Neither comfortable nor uncomfortable, Somewhat uncomfortable, Very uncomfortable | Determined using a self-developed item asking participants to rate, on a five-point Likert scale, how comfortable they would be with discussing preconception health and their pregnancy intentions with various people. These intervention delivery options were derived from suggestions and interventions identified in the literature as well as discussions within the study team. |
| Intervention delivery method last contact  (People/staff groups) | *Please select when you last spoke with each of the following people (about anything–not just about pregnancy)*  *(circle ONE of the following options for each person)*  Within the last week, Within the last month, Within the last year, 1-3 years ago, 4-5 years ago, More than 5 years ago, Never | Determined using a self-developed item asking participants to select, from seven options, when they last spoke to each of the people listed for the ‘Intervention delivery mode acceptability (People/staff groups)’ item. |
| **Variable** | **Question stem and response options** | **Item source, existing psychometric properties, and modifications made** |
| Intervention delivery method acceptability  (Places/ settings) | *Please rate how acceptable it would be to you if information about health before pregnancy was made available in the following places. Circle ONE letter for each place*  Very acceptable, Somewhat acceptable, Neither acceptable nor unacceptable, Somewhat unacceptable, Very unacceptable | Determined using a self-developed item asking participants to rate, on a five-point Likert scale, how acceptable they felt it would be for information about preconception health to be made available in a number of places or settings.  These intervention delivery options were derived from suggestions and interventions identified in the literature as well as discussions within the study team. |

**Additional file 2: Intervention delivery methods featured in the study questionnaire, with sources**

| **Intervention delivery method** | **Source(s)** |
| --- | --- |
| General practitioners (GPs)  Practice nurses  Obstetricians/gynaecologists  Midwives  Health visitors  Pharmacists  Dentists  Community/family support workers  Sexual health/family planning clinic staff  Hairdresser/beauticians  Friend(s)  Family/partners  Television  Billboards  Radio  Printed material in healthcare settings  On social media  By personal text or email (e.g. from a GP)  On preconception health websites/apps  Included with packs of tampons &sanitary pads  Included with pregnancy tests  Included in health education in schools  The workplace | (14) (15) (16) (17) (18)  (14) (15) (19) (20) (21) (22)  (14) (17) (23) (17)  (14) (19) (20)  (20)  (15) (24)  Study team  (16) (24)  (25) (15) (26)  (27)  (28)  (24)  (24) (29)  (27) (30)  (27) (30)  (22)  (31) (24)  (32)  (22)  (16)  (16)  (24) (16) (33) (29) (28) (34) (19)  (29) (24) |

**Additional file 3: Covariates included in each adjusted analysis**

i. ***Knowledge of preconception risk factors***

| **Outcome** | **Exposure** | **Variables adjusted for** |
| --- | --- | --- |
| Knowledge of each preconception risk factor | Age | Household income, Gravidity,  Pregnancy intentions, Infertility |
|  | Household income | Age, Education, Gravidity, Infertility |
|  | Educational attainment | Household income, Gravidity, Pregnancy intentions, Infertility |
|  | Ethnicity | Age, Education, Gravidity |
|  | Country of birth | Age, Education, Gravidity |
|  | Gravidity | Age, Household income, Education, Pregnancy intentions, Infertility |
|  | Livebirth(s) | Age, Household income, Education, Pregnancy intentions, Infertility |
|  | Adverse pregnancy outcomes | Age, Household income, Education, Pregnancy intentions, Infertility |
|  | Infertility | Age, Education, Gravidity |
|  | Pregnancy intentions | Age, Education, Gravidity |

ii. ***Attitudes towards preconception health***

| **Outcome** | **Exposure** | **Variables adjusted for** |
| --- | --- | --- |
| Perceived awareness of preconception risk factors | Age | Household income, Country of birth, Gravidity, Pregnancy desire, Infertility |
|  | Household income | Age, Gravidity |
|  | Educational attainment | Household income, Country of birth, Gravidity, Pregnancy desire, Infertility |
|  | Ethnicity | Age, Country of birth, Gravidity |
|  | Country of birth | Age, Gravidity, Pregnancy desire |
|  | Gravidity | Age, Household income, Country of birth, Pregnancy desire, Infertility |
|  | Livebirth(s) | Age, Household income, Country of birth, Pregnancy desire, Infertility |
|  | Adverse pregnancy outcomes | Age, Pregnancy desire, Infertility |
|  | Infertility | Age, Gravidity |
|  | Pregnancy intentions | Age, Country of birth, Gravidity |
| Perceived importance of preconception health | Age | Household income, Ethnicity |
|  | Household income | Age |
|  | Educational attainment | Age, Household income |
|  | Ethnicity | Age |
|  | Country of birth | Age, Ethnicity |
|  | Gravidity | Age, Household income, Ethnicity |
|  | Livebirth(s) | Age, Household income |
|  | Adverse pregnancy outcomes | Age |
|  | Infertility | Age |
|  | Pregnancy intentions | Age |
| **Outcome** | **Exposure** | **Variables adjusted for** |
| Interest in preconception health education | Age | Household income, Ethnicity, Gravidity, Pregnancy intentions |
|  | Household income | Age, Education, Gravidity |
|  | Educational attainment | Household income, Ethnicity, Gravidity, Pregnancy intentions |
|  | Ethnicity | Household income, Ethnicity, Gravidity, Pregnancy intentions |
|  | Country of birth | Age, Education, Gravidity |
|  | Gravidity | Age, Education, Ethnicity, Household income, Pregnancy intentions |
|  | Livebirth(s) | Age, Education, Household income, Pregnancy intentions |
|  | Adverse pregnancy outcomes | Age, Education, Gravidity, Pregnancy intentions |
|  | Infertility | Age, Education, Gravidity |
|  | Pregnancy intentions | Age, Education, Gravidity |
| Preconception health self-efficacy | Age | Household income, Education, Gravidity |
|  | Household income | Age, Education, Gravidity |
|  | Educational attainment | Household income, Country of birth, Gravidity |
|  | Ethnicity | Age, Education, Country of birth, Gravidity |
|  | Country of birth | Age, Education, Gravidity |
|  | Gravidity | Age, Household income, Education, Country of birth |
|  | Livebirth(s) | Age, Household income, Education, Country of birth |
|  | Adverse pregnancy outcomes | Age, Education |
|  | Infertility | Age, Education, Gravidity |
|  | Pregnancy intentions | Age, Education, Country of birth, Gravidity |
| Preconception lifestyle change intentions | Age | Ethnicity, Country of birth |
|  | Household income | *none* |
|  | Educational attainment | Ethnicity, Country of birth |
|  | Ethnicity | Country birth |
|  | Country of birth | Ethnicity |
|  | Gravidity | Ethnicity, Country of birth |
|  | Livebirth(s) | Country of birth |
|  | Adverse pregnancy outcomes | *none* |
|  | Infertility | *none* |
|  | Pregnancy intentions | Country of birth |

**Additional file 4: ‘Extraneous’ exposures* reported by participants (%), with accepted responses**

| **Preconception exposure** | **%** | **Accepted responses** |
| --- | --- | --- |
| Alcohol | 89.7 | Mentions alcohol or ’drinking’ as something to avoid/reduce/limit |
| Smoking | 89.3 | Mentions smoking, tobacco or second-hand smoke exposure as something to avoid/reduce/limit, or a smoke-free environment as something to aim for |
| Illicit drugs/substance abuse | 48.3 | Mentions (illegal/recreational) drugs or substance abuse as something to avoid/reduce/limit |
| Caffeine | 13.9 | Mentions caffeine, coffee or tea as something to avoid/reduce/limit |
| Avoidance of certain foods | 4.6 | Mentions the avoidance of certain foods (e.g. ‘certain meats’, ‘raw produce’) |
| Dieting | 1.9 | Mentions (crash) dieting or a restrictive diet as something to avoid |
| Hydration | 4.7 | Mentions remaining hydrated or drinking enough water as something to do/start/continue |
| Vitamins | 34.9 | Mentions (multi)vitamins, but does not explicitly mention that these should contain folic acid |
| Vitamin D | 1.8 | Mentions vitamin D as a positive |
| Vitamin B12 | 0.2 | Mentions vitamin B12 as a positive |
| Iron levels | 1.8 | Mentions iron levels or eating foods rich in iron as a positive |
| Omega 3 | 0.2 | Mentions Omega 3 supplementation |
| Hormones | 1.2 | Mentions hormonal (im)balance |
| Physical strain or injury | 10.7 | Mentions strenuous, excessive or extreme exercise, heavy lifting, travelling, ‘overdoing it’, ‘certain exercises’, or other activities involving risk of injury as something to avoid |
| General health and lifestyle | 7.8 | Mentions adopting a healthy ‘lifestyle’, optimising health, remaining healthy, ‘lifestyle changes’, general ‘social life’, ‘partying’, country versus city life, or having ‘unhealthy habits’ |
| Sleep | 16.0 | Mentions regular or sufficient sleep or rest as a positive, or shift work or ‘late nights’ as a negative |
| Stress | 51.3 | Mentions stress (including job stress), catastrophising, trying too hard to get pregnant, stressful life events, or an unstable lifestyle as a negative, or stress management as a positive |
| Self-care/Mental health | 33.1 | Mentions self-care, practising mindfulness, prioritising health or fun, aiming for a ‘balanced lifestyle’, ‘looking after’ oneself, relaxing, enjoying life, or resolving sources of conflict as a positive, or disregarding self-care (e.g. through ‘ignoring stress’) or comparing oneself to others as a negative  Or: Mentions mental health or wellbeing, emotional security or safety, having a ‘good mindset’, speaking to a therapist, or specific mental health considerations or conditions like trauma, post-traumatic stress disorder, anxiety and depression, or eating disorders |
| **Preconception exposure** | **%** | **Accepted responses** |
| Employment circumstances | 10.7 | Mentions job stability, employment terms (e.g. maternity pay), employer support or attitudes, work-life balance, or avoiding long hours or occupational exposure to chemicals or other sources of risk (e.g. cabin crew, manual handling) |
| Social support and relationships | 25.3 | Mentions support from family, friends or colleagues, having people to talk to, joining groups, investing in social relationships, the quality of or issues with these relationships, or avoiding negative people or influence  Or: Mentions the quality or stability of a romantic relationship, having problems with, receiving support from, or discussing concerns with a partner, relationship stress, or continuing to have frequent intercourse or a ‘healthy’ sex life |
| Multiple partners | 0.4 | Mentions having multiple sexual partners as something to avoid |
| Partner’s health | 1.6 | Mentions partner medical history, physical health, or health behaviours |
| Domestic environment | 15.1 | Mentions the home environment (e.g. whether it is clean, dry, safe), housing standards, living situation, environmental instability, or homelessness |
| Pollution | 4.2 | Mentions pollution, toxic substances, harmful chemicals, or fumes as something to avoid |
| Green space | 0.1 | Mentions access to green space |
| Immunisation | 2.2 | Mentions having vaccinations or checking for immunity to viruses |
| Infection risk | 1.1 | Mentions risk of viral infections such as COVID-19 (e.g. through living in an area with high prevalence) as a negative, or taking steps to reduce one’s risk of infection (e.g. through wearing a face covering or avoiding travel to countries with greater infectious disease risk) as a positive |
| Healthcare access and quality | 9.8 | Mentions accessing healthcare, taking advice from or seeing a general practitioner, pharmacist, obstetrician or midwife (e.g. for a check-up, medical tests, or a discussion about preconception health or one’s medical history), an individual’s relationship with these care providers, or the quality of healthcare they receive |
| Health conditions | 21.6 | Mentions physical health conditions or medical issues (e.g. their management), medical history, ‘bad health’, terminal illness or disease, or specific health conditions such as infection, pregestational hypertension, or diabetes |
| Sexual and reproductive health | 6.6 | Mentions sexual health, having a preconception sexual health check, sexually transmitted diseases, cervical screening, menstruation issues, female genital mutilation, or health conditions affecting the reproductive system such as polycystic ovary syndrome or endometriosis |
| Fertility | 1.9 | Mentions fertility issues, becoming informed of potential fertility issues, or having a fertility check |
| **Preconception exposure** | **%** | **Accepted responses** |
| Medications | 12.7 | Mentions prescription or over-the-counter medications, seeing a general practitioner to review current medications, or avoiding self-medication |
| Genetics | 10.3 | Mentions maternal genetics or familial medical history |
| Having/accessing appropriate information | 9.3 | Mentions accessing or seeking appropriate advice or information (e.g. to inform patient choice), having a good knowledge of pregnancy and childbirth and/or associated risks and risk factors, asking others about their experience of pregnancy, or avoiding unsubstantiated claims |
| Contraception | 4.0 | Mentions contraception as something to stop/avoid |
| Oral health | 0.7 | Mentions oral health, looking after one’s teeth, or seeing a dentist |
| Complementary therapy | 0.7 | Mentions complementary therapies such as reflexology, detoxification, or holistic therapy as a positive |
| Socio-economic status | 17.8 | Mentions (socio)economic or financial circumstances, income, being able to afford a pregnancy, or educational attainment |
| Pregnancy intendedness | 0.5 | Mentions pregnancy intendedness or the ‘circumstance of the way [a woman] got pregnant e.g. rape’ |
| Previous pregnancy outcomes | 2.9 | Mentions outcomes or ‘issues’ from previous pregnancies (e.g. miscarriage) |
| Breastfeeding | 0.1 | Mentions breastfeeding as something to stop before becoming pregnant again |
| Pelvic floor exercises | 2.4 | Mentions pelvic floor or Kegel exercises, or ‘maintaining pelvic health’ |
| Menstrual cycle | 2.5 | Mentions monitoring or ‘becoming familiar with’ one’s menstrual/ovulation cycle |
| Preparation for pregnancy/parenthood | 6.7 | Mentions financial (e.g. saving), domestic or mental preparation for pregnancy and associated changes, pregnancy ‘readiness’, ensuring things are ‘in place’, the ability to ‘plan ahead’, considering childcare arrangements, or taking parenting classes |
| Radiation | 0.8 | Mentions sources of radiation such as x-ray machines, mobile phones or microwaves as something to avoid |
| Hygiene | 0.5 | Mentions good hygiene as a positive |

Legend: *Preconception exposures listed by participants for which there was *no* high, moderate or low certainty evidence of an association(s) with an adverse pregnancy, birth or postpartum outcome(s) in Daly et al. (8)

| **Additional file 5: Proportion of participants who listed each preconception risk factor, by participant characteristic** | | | | | | | |
| --- | --- | --- | --- | --- | --- | --- | --- |
| **Characteristic** | **Preconception risk factor** | | | | | | |
|  | **Folic acid** | **Physical activity** | **Body mass index (BMI) / weight** | **Diet -Mediterranean/ high Alternate Healthy Eating Index** | **Abuse** | **Age** | **Interpregnancy intervals** |
| **Age**  18-24 years  25-29 years  30-34 years  35-39 years  40-48 years | 12.3 (7.5, 18.8)  21.5 (16.8, 26.8)  42.7 (34.1, 51.2)  53.2 (44.6, 61.7)  47.2 (38.9, 55.7) | 77.4 (69.7, 83.9)  75.6 (70.1, 80.6)  87.0 (80.0, 92.2)  80.6 (73.0, 86.8)  79.2 (71.6, 85.5) | 28.8 (21.6, 36.8)  38.9 (33.1, 44.9)  46.6 (37.8, 55.5)  42.4 (34.1, 51.1)  45.8 (37.5, 54.3) | 87.0 (80.4, 92.0)  84.7 (79.9, 88.8)  88.5 (81.8, 93.4)  85.6 (78.7, 91.0)  85.4 (78.6, 90.7) | 6.8 (3.3, 12.2)  3.6 (1.8, 6.6)  8.4 (4.3, 14.5)  9.4 (5.1, 15.5)  6.3 (2.9, 11.5) | 4.8 (1.9, 9.6)  6.2 (3.6, 9.7)  4.6 (1.7, 9.7)  7.9 (4.0, 13.7)   5.6 (2.4, 10.7) | 0.0 (0.0, 2.5†)  0.4 (0.0, 2.0)  0.8 (0.0, 4.2)  0.0 (0.0, 2.6)  0.0 (0.0, 2.5) |
| **Household income**  <£19,000  £19,000-£25,999  £26,000-£31,999  £32,000-£47,999  £48,000-£63,999  £64,000-£95,999  ≥£96,000 | 25.5 (17.5, 34.9)  23.5 (15.0, 34.0)  22.5 (13.5, 34.0)  30.3 (22.9, 38.5)  37.3 (29.8, 45.4)  40.4 (32.7, 48.4)  41.9 (31.8, 52.6) | 69.8 (60.1, 78.3)  72.9 (62.2, 82.0)  81.7 (70.7, 89.9)  79.6 (72.0, 85.9)  81.0 (74.0, 86.8)  83.2 (76.5, 88.6)  81.7 (72.4, 89.0) | 31.1 (22.5, 40.9)  28.2 (19.0, 39.0)  33.8 (23.0, 46.0)  37.3 (29.4, 45.8)  42.4 (34.6, 50.5)  45.3 (37.5, 53.3)  57.0 (46.3, 67.2) | 82.1 (73.4, 88.9)  85.9 (76.6, 92.5)  93.0 (84.3, 97.7)  85.9 (79.1, 91.2)  86.1 (79.7, 91.1)  86.3 (80.0, 91.2)  83.9 (74.8, 90.7) | 7.5 (3.3, 14.3)  8.2 (3.4, 16.2)  7.0 (2.3, 15.7)  4.9 (2.0, 9.9)  6.3 (3.1, 11.3)  6.2 (3.0, 11.1)  5.4 (1.8, 12.1) | 5.7 (2.1, 11.9)  7.1 (2.6, 14.7)  2.8 (0.3, 9.8)  4.2 (1.6, 9.0)  5.7 (2.6, 10.5)  6.8 (3.5, 11.9)   7.5 (3.1, 14.9) | 0.0 (0.0, 3.4)  0.0 (0.0, 4.2)  0.0 (0.0, 5.1)  0.0 (0.0, 2.6)  0.0 (0.0, 2.3)  0.6 (0.0, 3.4)  1.1 (0.0, 5.8) |
| **Ethnicity**  White  Minority ethnicity | 32.9 (29.5, 36.3)  34.9 (23.3, 48.0) | 79.4 (76.4, 82.2)  76.2 (63.8, 86.0) | 40.2 (36.7, 43.7)  36.5 (24.7, 49.6) | 85.9 (83.3, 88.3)  87.3 (76.5, 94.4) | 6.5 (4.9, 8.5)  3.2 (0.4, 11.0) | 6.1 (4.5, 8.1)  3.2 (0.4, 11.0) | 0.3 (0.0, 0.9)  0.0 (0.0, 5.7) |
| **Education**  School  Intermediate  University | 22.7 (13.3, 34.7)  27.0 (20.8, 34.0)  36.6 (32.6, 40.7) | 78.8 (67.0, 87.9)  70.3 (63.1, 76.8)  82.6 (79.2, 85.6) | 36.4 (24.9, 49.1)  41.6 (34.4, 49.1)  40.1 (36.0, 44.2) | 86.4 (75.7, 93.6)  83.8 (77.7, 88.8)  87.3 (84.3, 90.0) | 10.6 (4.4, 20.6)  5.9 (3.0, 10.4)  5.8 (4.0, 8.0) | 6.1 (1.7, 14.8)  5.4 (2.6, 9.7)  6.2 (4.3, 8.5) | 0.0 (0.0, 2.0)  0.4 (0.0, 1.3)  0.0 (0.0, 21.8) |
| **Country of birth**  United Kingdom  Other | 33.5 (30.1, 36.9)  28.4 (18.0, 40.7) | 78.8 (75.7, 81.6)  86.6 (76.0, 93.7) | 40.7 (37.2, 44.3)  31.3 (20.6, 43.8) | 85.5 (82.8, 87.9)  89.6 (79.7, 95.7) | 6.7 (5.0, 8.7)  3.0 (0.4, 10.4) | 5.4 (3.9, 7.3)  10.4 (4.3, 20.3) | 0.1 (0.0, 0.7)   1.5 (0.0, 8.0) |
| **Ever pregnant**  No  Yes | 19.8 (16.1, 23.9)  46.8 (41.8, 51.8) | 77.7 (73.4, 81.5)  80.7 (76.5, 84.4) | 36.7 (32.2, 41.5)  43.8 (38.9, 48.8) | 85.1 (81.4, 88.3)  86.9 (83.2, 90.0) | 4.4 (2.7, 6.8)  8.4 (5.9, 11.6) | 7.0 (4.8, 9.8)  4.7 (2.9, 7.2) | 0.2 (0.0, 1.3)  0.2 (0.0, 1.4) |
| **Livebirth(s)**  No  Yes | 22.4 (18.8, 26.2)  49.4 (43.8, 55.0) | 78.6 (74.8, 82.1)  79.9 (75.2, 84.2) | 36.5 (32.3, 40.8)  46.0 (40.5, 51.6) | 85.3 (81.9, 88.3)  87.0 (82.9, 90.5) | 4.7 (3.0, 6.9)  9.0 (6.1, 12.6) | 6.9 (4.8, 9.4)  4.3 (2.4, 7.1) | 0.2 (0.0, 1.1)  0.3 (0.0, 1.7) |
| **Characteristic** | **Preconception risk factor** | | | | | | |
|  | **Folic acid** | **Physical activity** | **Body mass index (BMI) / weight** | **Diet -Mediterranean/ high Alternate Healthy Eating Index** | **Abuse** | **Age** | **Interpregnancy intervals** |
| **Adverse outcomes**  No  Yes | 28.0 (24.6, 31.5)  57.1 (48.5, 65.5) | 79.7 (76.5, 82.6)  76.4 (68.5, 83.2) | 39.6 (36.0, 43.4)  42.9 (34.5, 51.5) | 86.0 (83.2, 88.5)  85.7 (78.8, 91.1) | 5.9 (4.3, 7.9)  8.6 (4.5, 14.5) | 6.2 (4.5, 8.3)  4.3 (1.6, 9.1) | 0.3 (0.0, 1.0)  0.0 (0.0, 2.6) |
| **Subfertility**  No  Yes | 30.8 (27.5, 34.4)  47.3 (37.7, 57.0) | 79.5 (76.4, 82.4)  77.3 (68.3, 84.7) | 39.0 (35.4, 42.7)  48.2 (38.6, 57.9) | 86.0 (83.3, 88.5)  86.4 (78.5, 92.2) | 6.2 (4.6, 8.2)  7.3 (3.2, 13.8) | 5.7 (4.1, 7.6)  7.3 (3.2, 13.8) | 0.3 (0.0, 1.0)  0.0 (0.0, 3.3) |
| **Pregnancy intentions**  Definitely not  Not sure/Don't know  In the next 3+ years  Within 2 years | 44.3 (37.8, 50.9)  27.3 (21.4, 33.8)  15.8 (10.9, 22.0)  42.8 (35.7, 50.1) | 78.3 (72.5, 83.4)  78.0 (71.8, 83.4)  78.1 (71.4, 84.0)  82.0 (75.8, 87.1) | 41.7 (35.3, 48.3)  38.8 (32.1, 45.7)  34.4 (27.6, 41.8)  44.8 (37.7, 52.1) | 83.8 (78.5, 88.3)  90.9 (86.2, 94.4)  86.9 (81.1, 91.4)  83.5 (77.5, 88.4) | 6.4 (3.6, 10.3)  10.5 (6.7, 15.5)  4.4 (1.9, 8.4)  4.1 (1.8, 8.0) | 6.4 (3.6, 10.3)  6.2 (3.4, 10.4)  4.9 (2.3, 9.1)  6.2 (3.2, 10.6) | 0.5 (0.0, 2.6)  0.0 (0.0, 2.0)  0.0 (0.0, 1.9)  0.0 (0.0, 2.3) |

**Additional file 6: Women’s attitudes towards preconception health**

| **Attitudinal variable** | **Response categories** | **N** | **% (95% CI)** |
| --- | --- | --- | --- |
| **Perceived awareness of preconception risk factors** | Not aware at all | 72 | 8.6 (6.9, 10.7) |
|  | Slightly aware | 244 | 29.2 (26.2, 32.4) |
|  | Moderately aware | 297 | 35.6 (32.4, 38.9) |
|  | Very aware | 222 | 26.6 (23.7, 29.7) |
|  | *Missing* | *0* |  |
| **Perceived importance of preconception health** | Strongly disagree | 5 | 0.6 (0.2, 1.4) |
|  | Disagree | 9 | 1.1 (0.6, 2.1) |
|  | Neither agree nor disagree | 47 | 5.6 (4.3, 7.4) |
|  | Agree | 412 | 49.3 (46.0, 52.7) |
|  | Strongly agree | 362 | 43.4 (40.0, 46.7) |
|  | *Missing* | *0* |  |
| **Interest in learning more about preconception health** | Not at all interested | 165 | 19.9 (17.3, 22.7) |
|  | Slightly interested | 208 | 25.0 (22.2, 28.1) |
|  | Moderately interested | 267 | 32.1 (29.0, 35.4) |
|  | Very interested | 191 | 23.0 (20.2, 26.0) |
|  | *Missing* | 4 |  |
| **Preconception health self-efficacy** | Strongly disagree | 3 | 0.4 (0.1, 1.1) |
|  | Disagree | 6 | 0.7 (0.3, 1.6) |
|  | Neither agree nor disagree | 63 | 7.6 (6.0, 9.6) |
|  | Agree | 383 | 46.1 (42.7, 49.5) |
|  | Strongly agree | 376 | 45.3 (41.9, 48.7) |
|  | *Missing* | 4 |  |
| **Preconception lifestyle change intentions** | Very unlikely | 13 | 1.6 (0.9, 2.7) |
|  | Quite unlikely | 43 | 5.2 (3.9, 6.9) |
|  | Neither likely nor unlikely | 67 | 8.1 (6.4, 10.1) |
|  | Likely | 313 | 37.7 (34.4, 41.0) |
|  | Very likely | 395 | 47.5 (44.1, 50.1) |
|  | *Missing* | *4* |  |
